# The Role and Impact of Self-Determination in Nursing Students: A Scoping Review

**DOI:** 10.1101/2025.09.16.25335504

**Authors:** Atikah Fatmawati, Anndy Prastya, Fitria Wahyu Ariyanti

## Abstract

**Background:** Self-determination is a crucial psychological aspect that impacts the motivation, academic performance, and overall well-being of nursing students. A comprehensive understanding of its influence and role can help nursing educators design more effective learning strategies.

**Purpose:** This scoping review aims to explore the influence and role of self-determination in nursing students and identify existing research gaps.

**Methods:** This review was conducted using the Arksey and O’Malley methodological framework, with guidance from the PRISMA-ScR guidelines. Articles were searched in the CINAHL, Scopus, and Proquest databases using the keywords “self-determination theory” OR “autonomous motivation” OR “self-determination”) AND (“nursing student*” OR “nursing undergraduate” OR “nursing education”) AND (“motivation” OR “academic engagement” OR “clinical competence”).

**Results:** Of the 83 articles found, 11 met the criteria. Analysis showed that self-determination plays a significant role in enhancing students’ intrinsic motivation, academic engagement, clinical competence, and psychological well-being. Supporting factors include autonomy support from faculty, a collaborative learning climate, and meaningful clinical experiences. However, research examining specific interventions to enhance self-determination is limited.

**Conclusion:** Self-determination plays a crucial role in supporting the academic and psychological success of nursing students. Further research is needed to develop effective intervention strategies that optimize self-determination in the context of nursing education.

## Introduction

Nursing education requires a combination of theoretical knowledge, clinical skills, and the ability to adapt to complex and dynamic practice situations. The success of this educational process is determined not only by structural factors (curriculum, facilities, clinical experiences), but also by student psychological factors, particularly motivation, clinical self-confidence, and psychological well-being, which collectively influence learning engagement, skill acquisition, and retention in the program (Nezhadhoseini et al., 2024; Biagioli et al., 2025).

One of the most influential theoretical frameworks for understanding motivation and behavioral regulation is Self-Determination Theory (SDT). SDT emphasizes three universal, basic psychological needs: autonomy (the need to feel in control of one’s actions), competence (the need to feel capable), and relatedness (the need to feel connected to others), which, when met, foster intrinsic motivation, adaptive performance, deep learning, and well-being (Hindman et al., 2025). In healthcare education settings, the fulfillment of these needs is often mediated by teaching practices (e.g., autonomy-supportive teaching), assessment design, and clinical dynamics between students, faculty, and preceptors.

Empirical evidence in nursing student populations demonstrates a consistent relationship between basic psychological need satisfaction and important educational outcomes such as academic engagement, academic motivation, self-efficacy, and performance. Several quantitative studies report that the level of need satisfaction is positively associated with learning engagement and motivation; others link autonomy-supportive teaching practices to increased internal motivation and clinical confidence in students. These findings highlight the role of SDT as a relevant theoretical foundation for understanding how learning contexts (including assessments, simulation practice, and clinical interactions) influence nursing student outcomes (Chen & Zhang, 2022; Davies, 2024).

Instrumentally, healthcare education research often uses several tools to measure SDT constructs and academic motivation. The most commonly used instruments include the Academic Motivation Scale (AMS) to measure motivational dimensions (intrinsic, extrinsic, and amotivation), and the Basic Psychological Need Satisfaction and Frustation Scale (BPNSFS). For the specific nursing population, specific tools such as the Motivation for Nursing Students Scale (MNSS) have been developed and validated, demonstrating adequate psychometric properties in large samples of nursing students. These instruments allow for more focused quantitative studies on the relationship between SDT-related constructs and nursing educational outcomes (Souza et al., 2021; Bulfone et al., 2021).

On the intervention side, the literature also suggests that autonomy-supportive learning settings (e.g., learning that provides meaningful choices, provides feedback that supports competency, and builds supportive relationships) have the potential to enhance students’ autonomous motivation and learning outcomes. However, the influence of aspects such as assessment type (especially high-value assessments), learning format (online vs. offline), and preceptorship characteristics on student psychological needs and outcomes still requires more systematic evidence synthesis. Reviews in health professions education emphasize that assessments can significantly influence motivation, sometimes dampening autonomous motivation if assessment design is not carefully designed. Therefore, curriculum design and assessment strategies need to be considered within the SDT framework (Hamm et al., 2024; Kursurkar et al., 2023).

While there are numerous cross-sectional studies and several interventions in health education contexts, the evidence is scattered across designs, geographic settings, measurement instruments, and outcomes. Some studies only assess motivation in general (e.g., AMS), while others explore specific aspects such as clinical skills self-efficacy, practice satisfaction, or program retention intentions. Design variability and outcome heterogeneity make it difficult to draw general conclusions about the magnitude and direction of the influence of self-determination on nursing education outcomes. Furthermore, there is a need to assess the methodological quality of existing studies and clarify the mechanisms (e.g., whether autonomy fulfillment is more important for engagement while competence is more related to clinical performance) (Rafii et al., 2019; Chen & Zhang, 2022).

With this background, a systematic synthesis of existing evidence is needed to summarize the strength of the evidence regarding the influence and role of self-determination constructs (autonomy, competence, relatedness) on key nursing student outcomes (academic motivation, engagement, clinical competence, academic performance, and well-being); identify effective teaching practices or interventions to support students’ psychological and motivational needs; and assess the methodological quality of studies and formulate research and practice recommendations. Such a review will assist nursing educators, curriculum designers, and educational evaluators in designing learning environments that better support adaptive motivation, enhance clinical skill acquisition, and minimize the risk of student burnout/turnover (Davies, 2024).

Based on this gap, this study will conduct a scoping review aimed at summarizing and evaluating empirical evidence on the influence and role of self-determination in nursing students, identifying the instruments used, assessing the methodological quality of the studies, and suggesting practical implications for nursing education and future research agendas.

## Method

### Data Retrieval Strategy

This study is a scoping review to systematically answer the research questions. It is structured according to the Preferred Reporting Items for Systematic Reviews and Meta-Analyses (PRISMA) 2020 guidelines (Page et al., 2021). This study reviews previous research on self-determination in nursing students. The detailed activities include determining data search strategies and/or information sources, selecting studies through quality assessment based on eligibility criteria and quality assessment instruments, and data synthesis and extraction.

This scoping review addresses two research questions: 1) What has been published about the influence and role of self-determination on nursing student learning outcomes and well-being in various nursing education contexts? 2) What are the most frequently studied outcomes related to self-determination in nursing students, such as academic motivation, engagement, academic achievement, clinical competence, self-efficacy, retention intentions, and psychological well-being (stress, burnout, thriving)? 3) What are the most frequently studied outcomes related to self-determination in nursing students, such as academic motivation, engagement, academic achievement, clinical competence, self-efficacy, retention intentions, and psychological well-being (stress, burnout, thriving)?

The literature search was conducted in three international databases: CINAHL (Ebsco), Scopus, and ProQuest. The keywords and Boolean operators used in the literature search were “self-determination theory” OR “autonomous motivation” OR “self-determination” AND “nursing student” OR “nursing undergraduate” OR “nursing education” AND “motivation” OR “academic engagement” OR “clinical competence”.

### Eligibility Criteria

Eligibility criteria in this study included inclusion and exclusion criteria. The inclusion criteria were nursing student populations, quantitative, qualitative, or mixed-method studies, measuring or evaluating aspects of self-determination, full-text articles, and peer-reviewed studies. Exclusion criteria included editorials, commentaries, or non-research studies, non-nursing populations, and studies lacking primary data. Furthermore, to limit the scope of the study, the researchers used the PCC (Population, Concept, Context) method, as shown in the following table:

**Table 1.** PCC Summary.

| Component | Information |
| --- | --- |
| *Population (P)* | *"nursing student", undergraduate/associate/BSN/diploma/professional nurse students* |
| *Concept (C)* | *"self-determination theory", "autonomous motivation", "autonomy support", "basic psychological needs", "competence", "relatedness"* |
| *Context (C)* | *motivation, engagement, academic performance, clinical competence, self-efficacy, well-being/burnout, retention/intention to persist* |

### Quality Assessment

Literature selection used the PRISMA (Preferred Reporting Items for Systematic Reviews and Meta-analyses) method. The PRISMA Flow Diagram in this study is shown in Figure 1.

**Figure 1.**
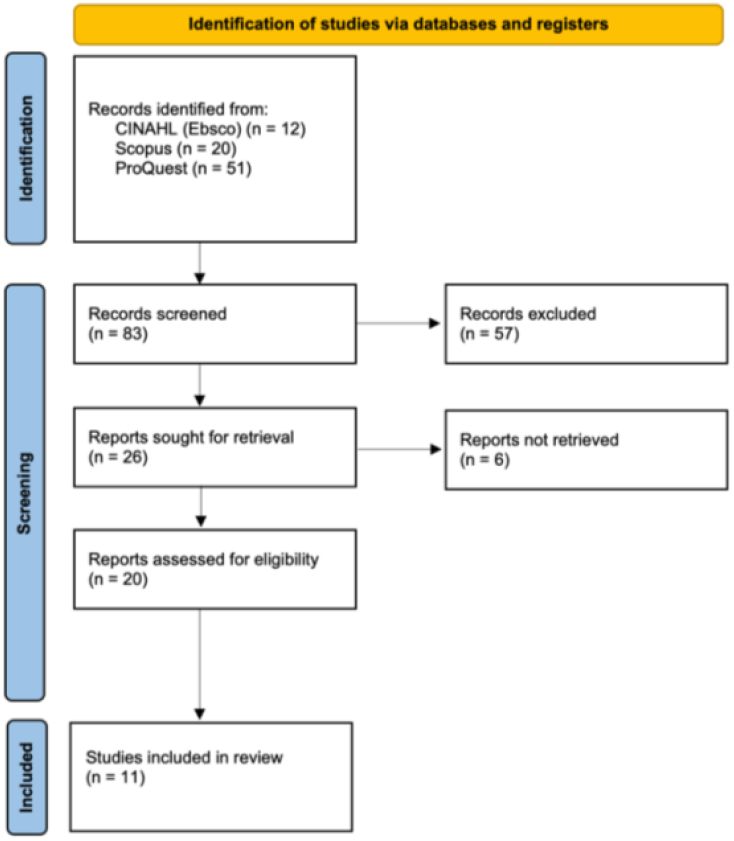
PRISMA *Flow Diagram*.

Based on the PRISMA method, 57 of the 83 journal articles identified were excluded due to data duplication, and 6 others did not meet the inclusion and exclusion criteria. Consequently, only 11 journal articles were used in the literature review.

### Synthesis Data

The data synthesis process in this study was conducted by comparing literature that met quality assessments and inclusion and exclusion criteria. The data synthesis was based on the research objective of analyzing the influence and role of self-determination in nursing students.

### Data Extraction

The data extraction output is in the form of a table consisting of the title, researcher’s name and year of publication, study design, participants, Self-Determination Theory (SDT) focus, and main findings.

## Results

Based on the selection process, the research using a scoping review technique on the influence and role of self-determination in nursing students identified 11 articles for further analysis. A summary of the article analysis results on the influence and role of self-determination in nursing students is outlined in Table 2.

**Table 2.**
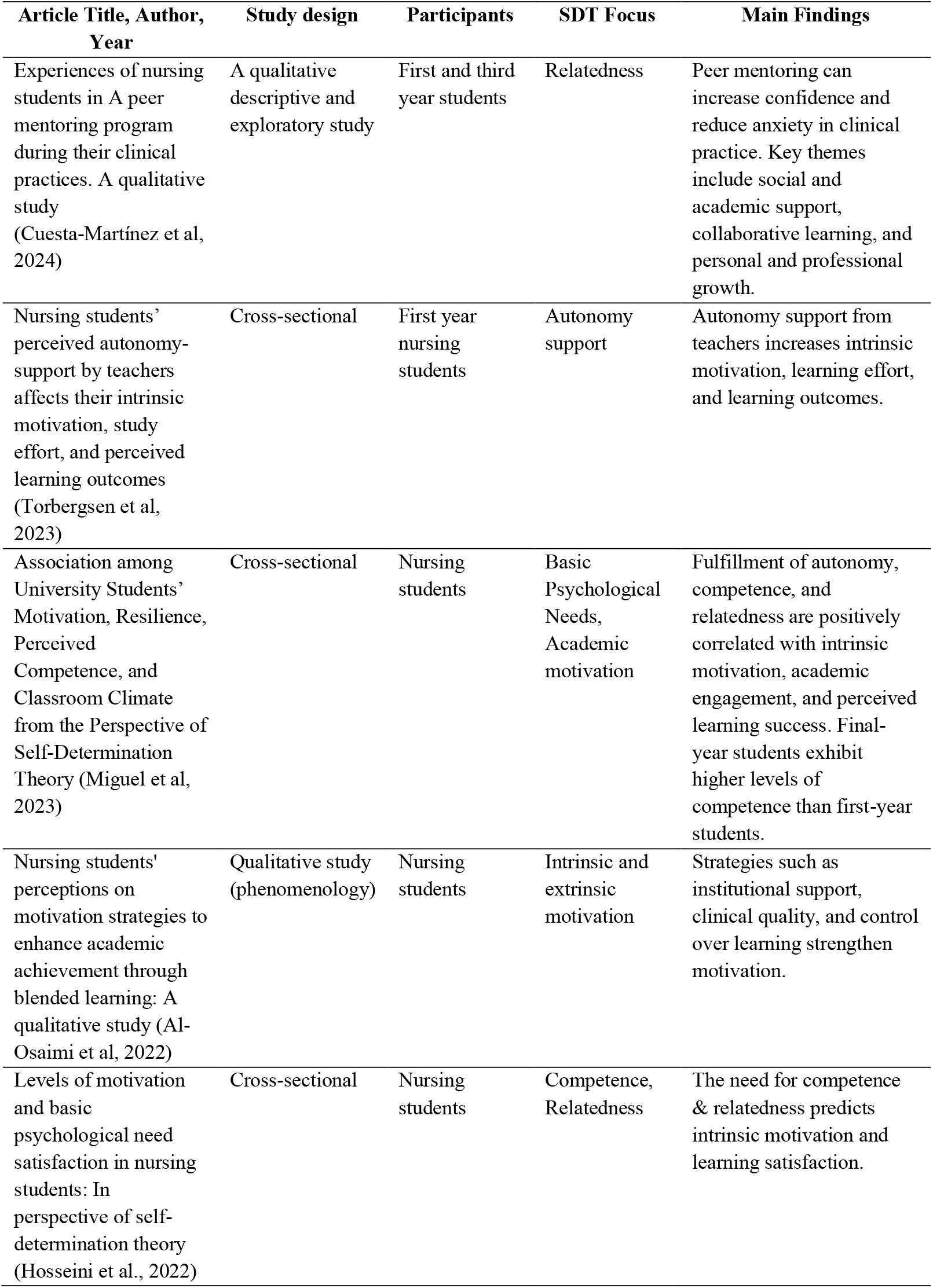
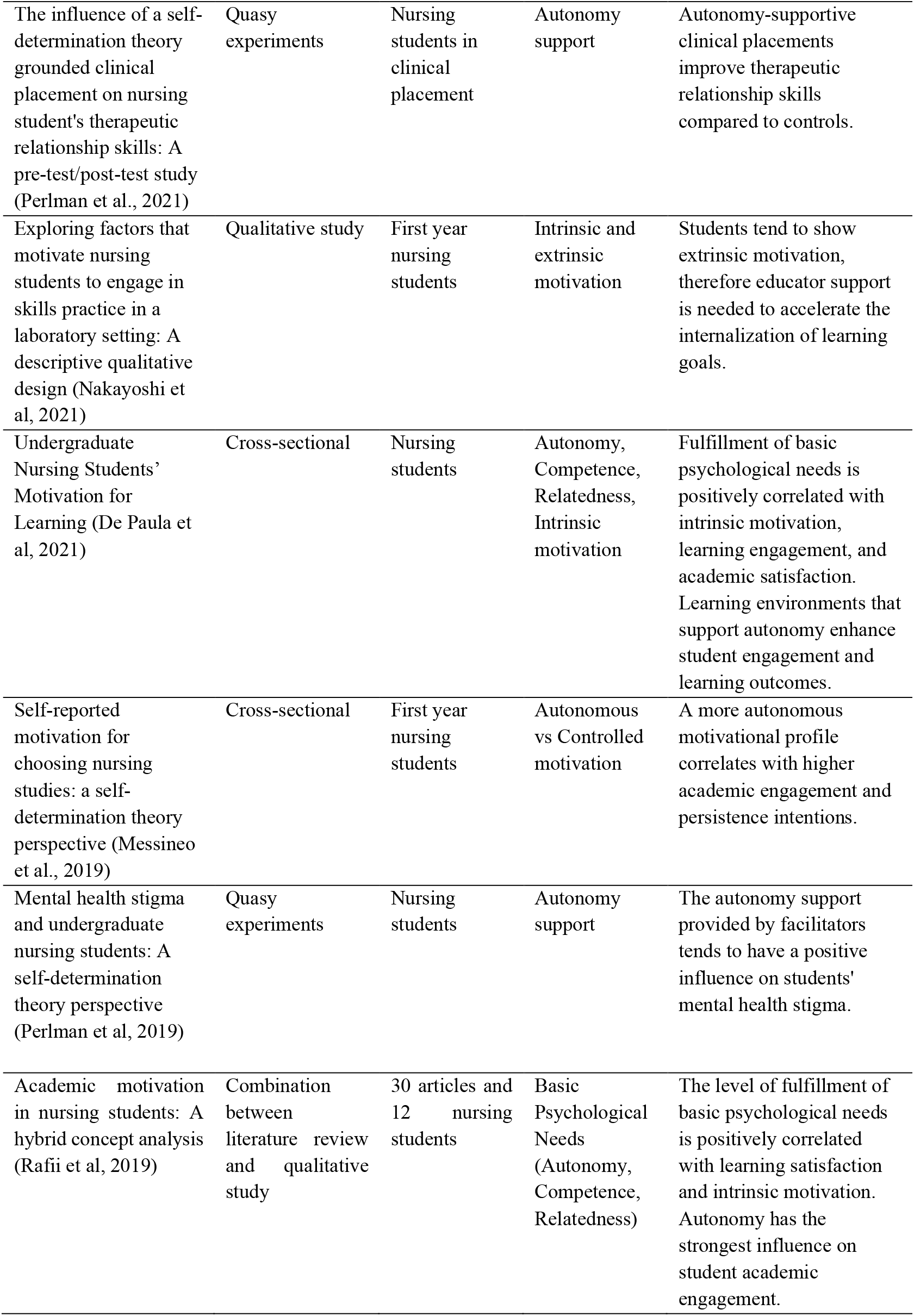
Data Extraction Results.

Table 2 shows the 11 articles analyzed in this scoping review. The research designs varied, including literature reviews, cross-sectional studies, quasi-experimental studies, qualitative studies, and scoping reviews. All of the reviewed studies involved nursing students from various levels in several countries.

## Discussion

Self-Determination Theory (SDT), developed by Deci and Ryan, explains that human motivation is influenced by three basic psychological needs: autonomy (independent decision-making), competence (feelings of ability and effectiveness), and relatedness (positive social connectedness) (Deci & Ryan, 2000). The results of this scoping review confirm that self-determination has a significant influence on motivation, academic engagement, clinical competence, and the psychological well-being of nursing students. In the context of nursing education, this theory serves as an important framework for understanding how students’ intrinsic motivation is formed and influences the learning process, clinical performance, and professional development. Fulfilling these basic psychological needs has been shown to increase intrinsic motivation and learning behaviors that are oriented toward mastery, rather than simply achieving grades.

Hamm & Yeh (2024) conducted a scoping review to identify factors influencing the academic motivation of undergraduate nursing students. Using the PRISMA-ScR (Preferred Reporting Items for Systematic Reviews and Meta-Analyses extension for Scoping Reviews) approach, they searched literature from databases such as CINAHL, ERIC, Education Full Text, ProQuest, and PubMed, with publications ranging from 2019 to 2024. Of the 461 articles screened, only 10 studies met the inclusion criteria. The types of studies found were generally descriptive, cross-sectional, or correlational.

The analysis of these studies revealed four main themes as determinants of nursing students’ academic motivation: autonomy, competence, social support, and coping (stress management strategies). Through this review, Hamm and Yeh concluded that academic motivation is positively correlated with these four factors: students who feel more autonomous, competent, have strong social support, and can cope well with academic pressure tend to demonstrate higher motivation.

Torbergsen (2023) investigated how nursing students’ perceived autonomy support from their instructors impacted their intrinsic motivation, learning effort, and perceived learning outcomes. The study was conducted at a large Norwegian university using a flipped classroom approach, focusing on Cardiopulmonary Resuscitation (CPR) education. Cross-sectional data were collected from 391 first-year nursing students between 2018 and 2021, using instruments such as the Learning Climate Questionnaire (LCQ), the Intrinsic Motivation Inventory (IMI), and a perceived learning outcomes scale.

The results showed that students’ perceived autonomy support from their instructors directly and significantly increased their intrinsic motivation and their perceptions of their learning outcomes (De Paula et al., 2021; Miguel et al., 2023). Intrinsic motivation, in turn, indirectly increased students’ learning effort, and the impact of autonomy support also had an indirect effect on learning effort through increased intrinsic motivation (Nakayoshi et al., 2021). In other words, instructors who create a learning environment that supports autonomy, for example, by providing choices, rational explanations, and space for collaboration, can directly increase students’ intrinsic motivation and perceived learning outcomes, while simultaneously encouraging them to be more engaged in their studies (Perlman et al., 2019; Al-Osaimi et al., 2022).

These results align with the principles of Self-Determination Theory, particularly regarding the importance of psychological needs in fostering intrinsic motivation. Autonomy is a basic need, fulfilled when instructors provide space for students to choose and take initiative, while competence and relatedness also play a crucial role (Rafii et al., 2019). Research shows that nursing students with high levels of autonomous motivation tend to have better learning engagement, resilience in the face of academic challenges, and optimal psychological well-being (Messineo et al., 2019; Hosseini et al., 2022). Satisfaction of the need for competence and relatedness has also been shown to be an important predictor of intrinsic motivation in nursing students, thus supporting optimal academic and clinical achievement (Pishkar et al., 2022; Cuesta-Martínez et al., 2024).

The application of SDT has also been shown to be significant in clinical practice environments. Clinical learning environments that support student autonomy, for example, through the freedom to make clinical decisions under supervision, have been shown to improve therapeutic communication skills, self-confidence, and reduce stigma toward patients, particularly in the area of mental health nursing practice (Perlman et al., 2019; 2021; 2023). Conversely, students with more controlled motivation tend to show decreased confidence and therapeutic skills when faced with complex clinical situations.

In the context of clinical practice, self-determination has been shown to influence students’ professional attitudes. A Japanese study found that students with high levels of self-determination demonstrated more empathy and less stigmatization toward patients with mental illness after participating in clinical practice compared to students with low levels of self-determination (Mizuno et al., 2019). These results confirm that autonomous motivation not only supports cognitive competence but also develops affective qualities important in nursing. The positive effects of self-determination are also evident in clinical skill mastery. A study by Ebrahimi et al. (2023) showed that students with high levels of autonomous motivation during mental health clinical placements exhibited better therapeutic skills and self-confidence than those with moderate or low levels of motivation. This reflects the evidence that student-centered and autonomy-supportive learning can optimize professional readiness.

In addition to influencing the learning process, SDT also plays a role in shaping behaviors and decision-making that support a healthy lifestyle and professional readiness in nursing students (Shin et al., 2013). Therefore, implementing SDT principles in the curriculum and clinical practice supports not only academic success but also the ongoing development of character and professional competence.

Based on these findings, nursing educational institutions are recommended to design learning strategies and clinical supervision that focus on meeting students’ three primary psychological needs. This approach can be implemented by providing autonomy in decision-making, offering constructive feedback to enhance competency, and fostering supportive mentor-student relationships. These efforts are believed to improve the quality of learning, student retention, and their readiness to become competent professional nurses oriented toward empathy-based practice.

## Conclusion

Overall, these findings underscore the importance of integrating SDT-based approaches into curriculum design and teaching methods. Practical strategies that can be implemented include providing autonomy in learning decision-making, providing meaningful clinical experiences, and strengthening social support in the academic environment. Future research focused on SDT-based interventions is needed to strengthen empirical evidence. These interventions include experiential learning, academic coaching, or the use of digital technology to strengthen students’ intrinsic motivation. With this approach, the quality of learning and professional preparedness of nursing students can significantly improve.

## Data Availability

All data produced in the present study are available upon reasonable request to the authors.

